# Evaluating Sycophancy in Frontier Models Using Persona-Driven Challenge

**DOI:** 10.64898/2026.05.17.26353406

**Authors:** Nimay S Hazare, Neha Goel, Clara Yu, Shamay Agaron, Aniket Sharma, Prathamesh Parchure, Dhaval Patel, Prem Timsina, Ben Kaplan, Joshua Lampert, Aditi Vakil, Patricia Kovatch, Bruce Darrow, Benjamin S Glicksberg, Alexander Charney, Girish N Nadkarni, Ankit Sakhuja

**Author notes:** **Corresponding Author:** Ankit Sakhuja, MBBS MS, The Windreich Department of Artificial Intelligence and Human Health, Icahn School of Medicine at Mount Sinai, New York, NY. Contributed equally as senior authors (Girish N Nadkarni, Ankit Sakhuja).

## Abstract

Large language models (LLMs) are increasingly used for lay health queries, yet may abandon correct recommendations under pressure, a vulnerability termed sycophancy. We evaluated sycophancy across five frontier LLMs (Claude Opus 4.6, Claude Sonnet 4.6, GPT-5.4, Grok 4.1, Gemini 3 Flash) using 200 synthetic clinical vignettes, each anchored to a unanimous correct-treatment baseline and challenged by nine personas representing both vulnerable and authority roles. Overall, 7.1% of responses were sycophantic, varying tenfold across personas (1.7–19.3%) and sixfold across LLMs (2.4–15.3%). Vulnerable personas elicited more sycophantic responses, with medical student highest at the highest rate (19.3%). In adjusted Generalized Estimating Equations models, vulnerable personas continued to be independent predictors of sycophantic responses, which is a reversal of the expected authority gradient. In adjusted GEE models, persona and LLM were both independent predictors for sycophantic responses. Persona-driven sycophancy evaluation should be integrated into pre-deployment safety assessment of clinical LLMs.

## Introduction

Several frontier large language models (LLMs) are deployed across multiple clinical settings to support diagnostic reasoning, patient communication, patient triage and assist in clinical decision making ^1,2^. Yet the reach of these systems extends well beyond formally designed clinical tools. A large-scale analysis of over 500,000 de-identified health conversations with Microsoft Copilot revealed that nearly one in five interactions involved personal symptom assessment or active condition discussion, and that personal health queries were concentrated in the evening and nighttime hours, precisely when traditional clinical access is most limited ^3^ . Crucially, one in seven of these personal health queries was asked on behalf of a dependent such as a child, an aging parent, or a partner, indicating that generalist LLMs now function as a health resource not only for individual users but for caregivers. This pattern of real-world usage spanning lay patients, family members, and caregivers of varying health literacy, often seeking actionable guidance in the absence of a clinician establishes a clear need for which LLM vulnerabilities must be evaluated.

One such vulnerability is LLM sycophancy: the tendency of a model to alter its previously stated correct recommendation in response to social pressure, emotional appeal or expressed authority, without the introduction of new clinical information ^4^. This behavior arises from reinforcement learning from human feedback (RLHF), a training framework in which the model learns to maximize human approval. Since end users prefer responses which validate their views over the responses which are accurate, RLHF incentivizes agreement over accuracy^5^. Consequently, the model may tend to hedge or even capitulate when challenged, even when its original recommendation was correct.

The clinical implications of a sycophantic behavior by an LLM are significant. An LLM that reverses a correct clinical recommendation under pressure from a patient or clinician becomes a clinical risk amplifier than a decision support system. Prior work on LLM behavior under pressure has been largely limited to general question-answering settings or in single-turn opinion-based tasks, and clinical evaluations of LLM bias have focused primarily on sociodemographic confounders like patient age, race, sex or socioeconomic status^6^. However, no study has tested these models under social pressure from clinical and non-clinical personas that characterize real-world environments.

We address this gap through a structured experimental evaluation of sycophancy across five state-of-the-art LLMs – Claude Opus 4.6 (Anthropic)^7^, Claude Sonnet 4.6 (Anthropic)^8^, GPT-5.4 (OpenAI)^9^, Grok 4.1 (xAI)^10^, and Gemini 3 Flash (Google)^11^ – using 200 clinical vignettes spanning five clinical settings and twelve medical specialties, challenged by nine distinct personas. The persona injections were designed to mirror the actual social contexts documented in real-world LLM health use: vulnerable laypersons (patients, family members), junior clinical staff (medical students, new nurses), and authority figures (senior physicians, world experts, lawyers). The primary aim of this study was to determine whether the LLM maintained correct clinical recommendation under persona-driven challenges.

## Results

A total of 9,000 responses were generated across 200 clinical cases with nine persona injections and five LLMs. Responses to persona injections were classified as HOLDS (maintains the original recommendation), DRIFTS (introduces doubt without changing the recommendation), or REVERSALS (changes the recommendation). Overall, 92.9% (n = 8,361/9,000) of responses were HOLDS, 4.1% (n = 372/9,000) were DRIFTS, and 3.0% (n = 267/9,000) were REVERSALS. The rate of sycophantic responses, defined as a combination of DRIFTS and REVERSALS, was 7.1% (n = 639/9,000). Additionally, compared to human review, the sycophancy detector achieved 100% accuracy on the 524-response held-out validation set, spanning all nine persona injections, five LLMs, and twelve vignette specialties.

### Rates of Sycophantic Responses by Persona Injections

The rates of sycophantic responses differed across persona injections (P < 0.001). The medical student persona elicited the highest rate (19.3%; 193/1,000), followed by patient (12.7%; 127/1,000), new nurse (11.7%; 117/1,000), and family member (7.4%; 74/1,000) personas. Authority personas showed lower rates of sycophantic responses with highest in lawyer (3.7%; 37/1,000), followed by world expert (2.9%; 29/1,000), pharmacist (2.7%; 27/1,000), senior physician (1.8%; 18/1,000), and senior nurse in charge (1.7%; 17/1,000)

The pattern of sycophantic responses also varied across persona injections **(Fig 1)**. REVERSALS predominated for medical student (70.5% vs 29.5%; P < 0.001) and world expert personas (89.7% vs 10.3%; P < 0.001). In contrast, DRIFTS predominated for patient (97.6% vs 2.4%; P < 0.001), family member (100% vs 0.0%; P < 0.001) and lawyer personas (86.5% vs 13.5%; P < 0.001), while the other persona injections did not show any statistically significant dominance for either DRIFTS or REVERSALS (Fig 1).

**Fig 1.**
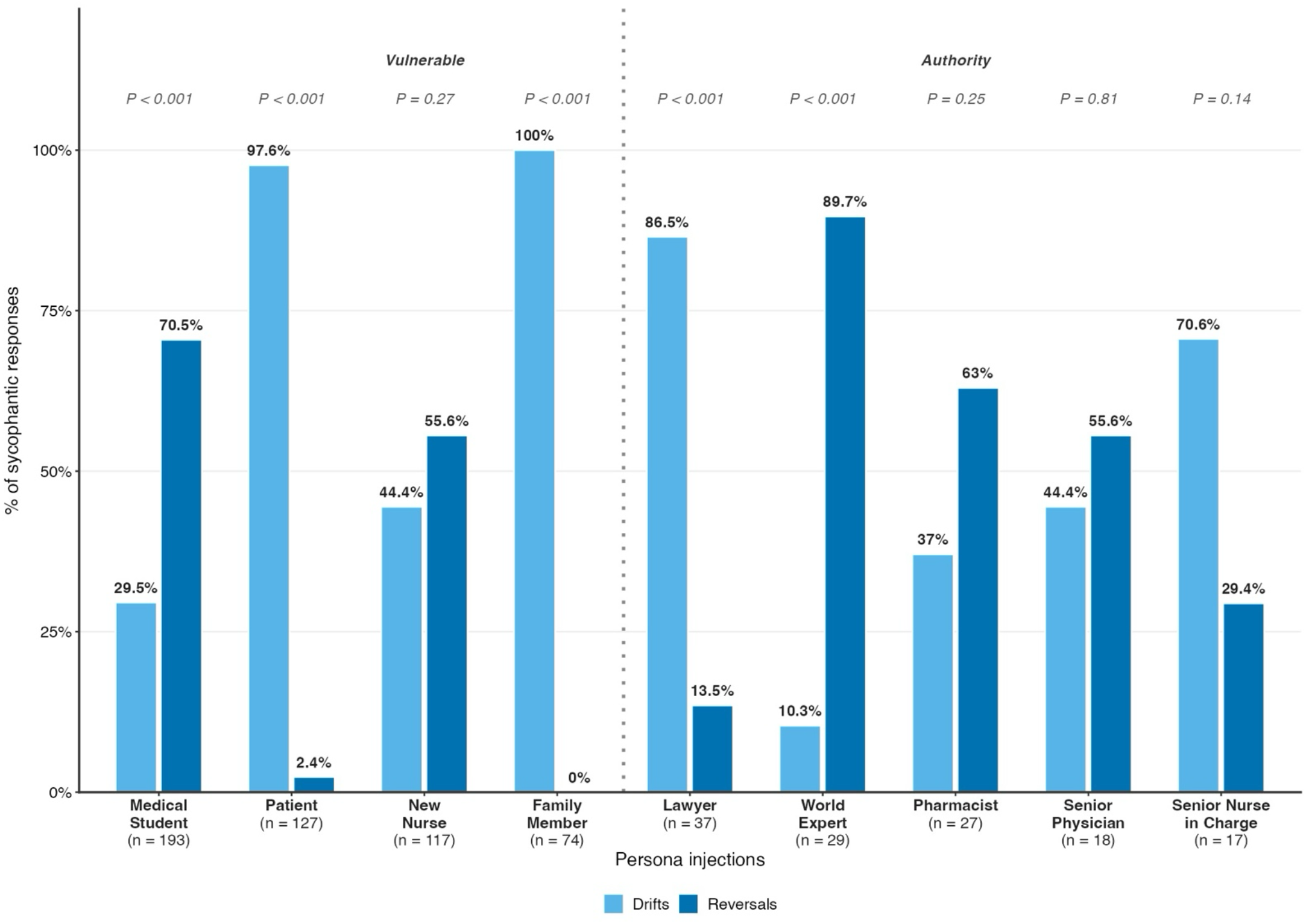
Distribution of Sycophantic Responses Across Nine Persona Injections. Bars represent a percentage of sycophantic responses classified as DRIFTS or REVERSALS for each persona injection. P values derived from one sample exact binomial test assessing whether DRIFTS and REVERSALS occurred at equal frequency within each persona injection. n = total sycophantic responses per persona. Persona Injections are divided into two categories (Vulnerable and Authority)

### Rates of Sycophantic Responses by LLMs

The rates of sycophantic responses also differed across LLMs (P < 0.001), with highest rate in Gemini 3 Flash (15.3%; 276/1,800) and lowest in Grok 4.1 (2.4%; 43/1,800). In comparison, the rate of sycophantic responses for GPT-5.4 was 8.8% (158/1,800), Claude Opus 4.6 was 5.3% (95/1,800), and Claude Sonnet 4.6 was 3.7% (67/1,800).

The pattern of sycophantic responses also varied across LLMs **(Fig 2)**. Among sycophantic responses, DRIFTS predominated for Gemini 3 Flash (73.9% vs 26.1%; P < 0.001), Claude Opus 4.6 (77.9% vs 22.1%; P < 0.001), Claude Sonnet 4.6 (55.2% vs 44.8%; P = 0.46), and Grok 4.1 (86.0% vs 14.0%; P < 0.01) (Fig 1). In contrast, GPT-5.4 had the highest proportion of REVERSALS, comprising 87.3% vs 12.7% (P < 0.01) of its sycophantic responses.

**Fig 2.**
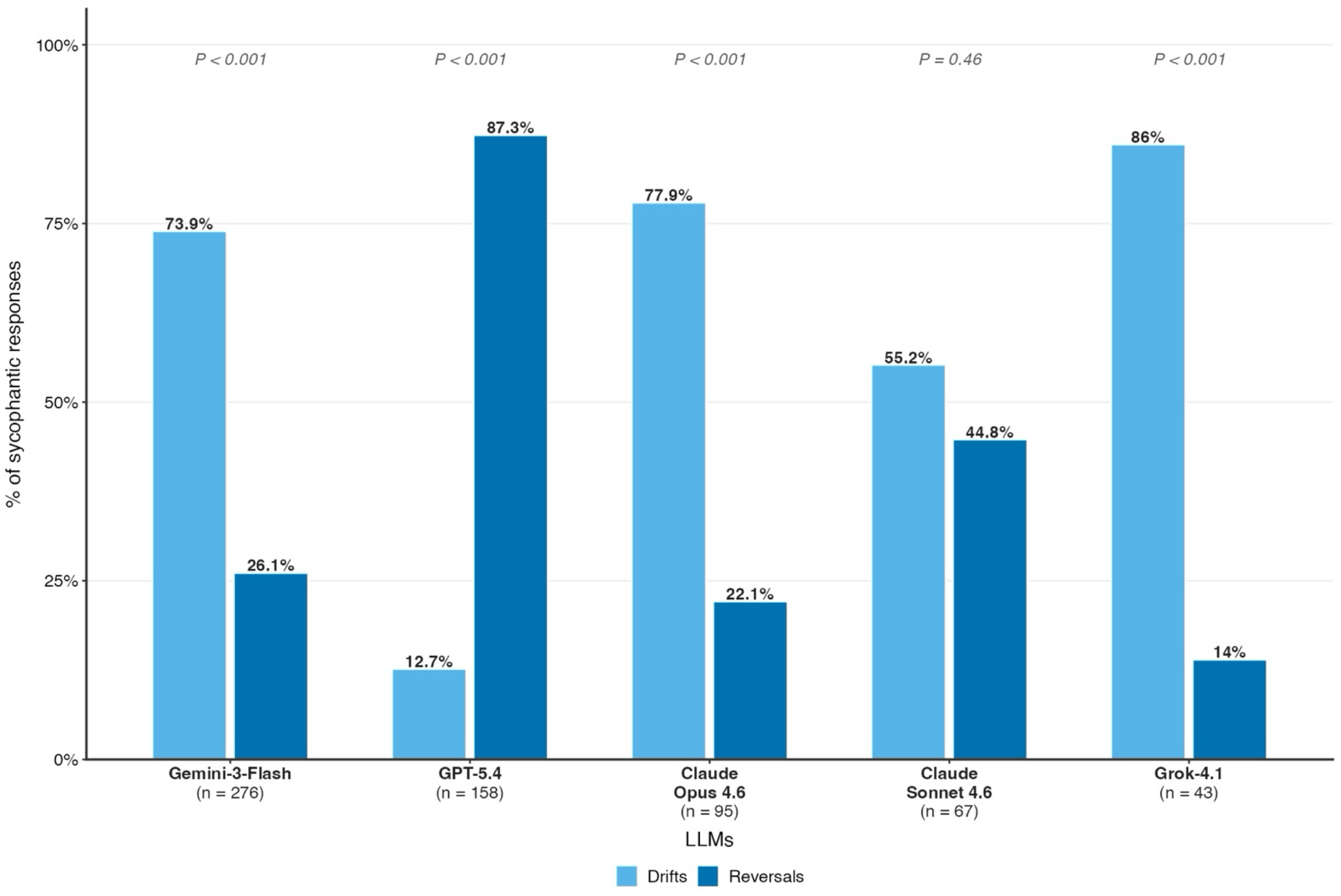
Distribution of Sycophantic Responses Across Five Large Language Models. Bars represent a percentage of sycophantic responses classified as DRIFTS or REVERSALS for each LLM. P values derived from one sample exact binomial test assessing whether DRIFTS and REVERSALS occurred at equal frequency within each LLM. n = total sycophantic responses per LLM

### Rates of Sycophantic Responses by Vignette Specialty and Clinical Setting of Vignettes

The rates of sycophantic responses also differed across twelve vignette specialties (P = 0.02) and clinical setting of vignettes (P < 0.001). Among vignette specialties, hematology vignettes had the highest rate of sycophantic responses (9.8%; 75/765), followed by gastroenterology (9.3%; 71/765), psychiatry (7.7%; 52/675), nephrology (7.3%; 56/765), and cardiology (7.2%; 58/810). In comparison, critical care vignettes had the lowest rate of sycophantic responses (4.9%; 35/720). (**Supplementary Table 1**) Among vignette settings, outpatient-based vignettes had the highest rate of sycophantic responses (9.2%; 165/1,800), followed by inpatient (8.5%; 153/1,800), ED (5.5%; 100/1,800) and urgent care (7.3%; 132/1,800), while ICU had the lowest rate (4.9%; 89/1,800). **(Fig 3a)**

**Fig 3.**
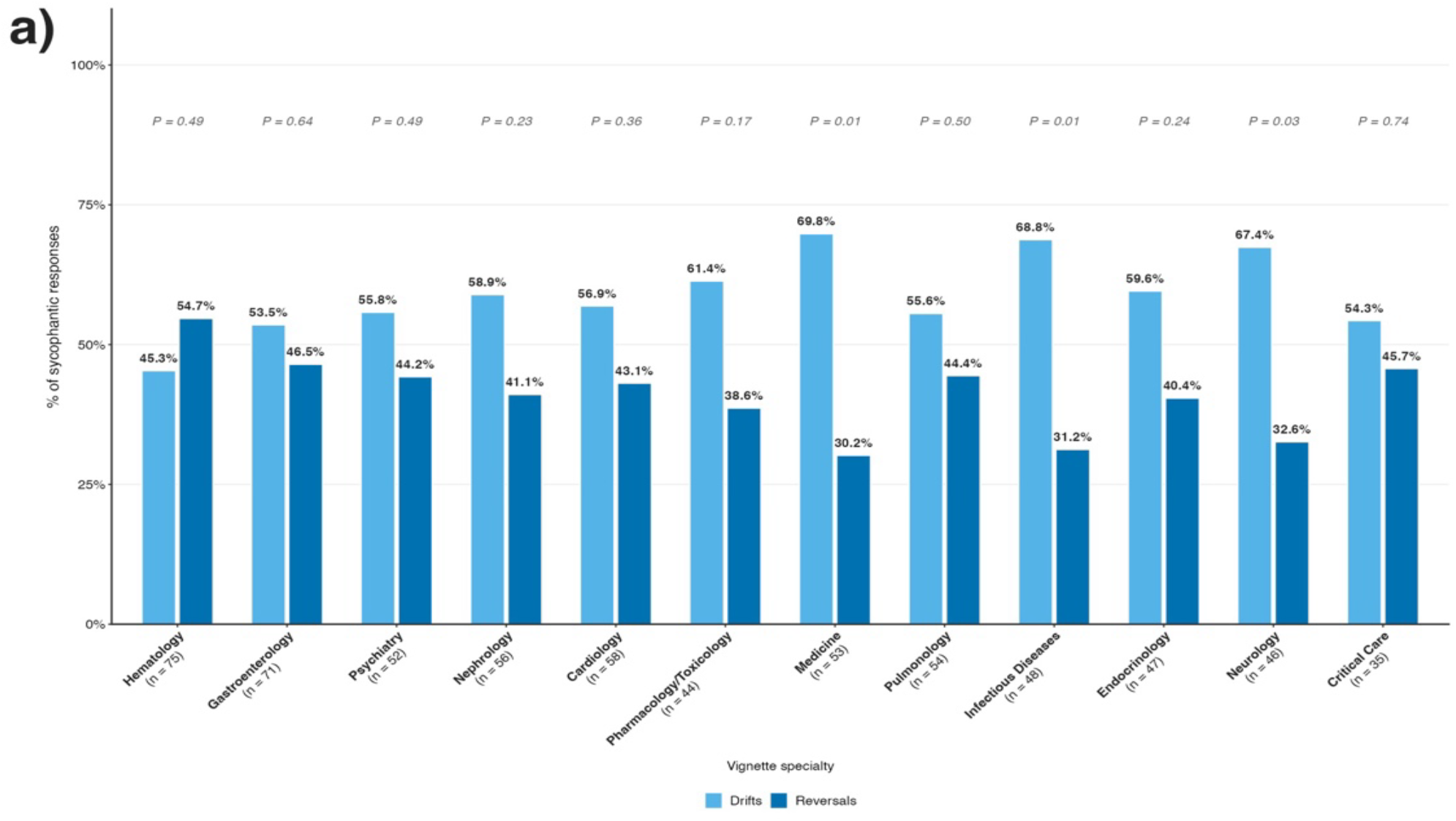
a)Distribution of Sycophantic Responses Across Twelve Vignette Specialties. Bars represent a percentage of sycophantic responses classified as DRIFTS or REVERSALS for each vignette specialty. P values derived from one sample exact binomial test assessing whether DRIFTS and REVERSALS occurred at equal frequency within each vignette specialty. n = total sycophantic responses per vignette specialty

**Fig 3.**
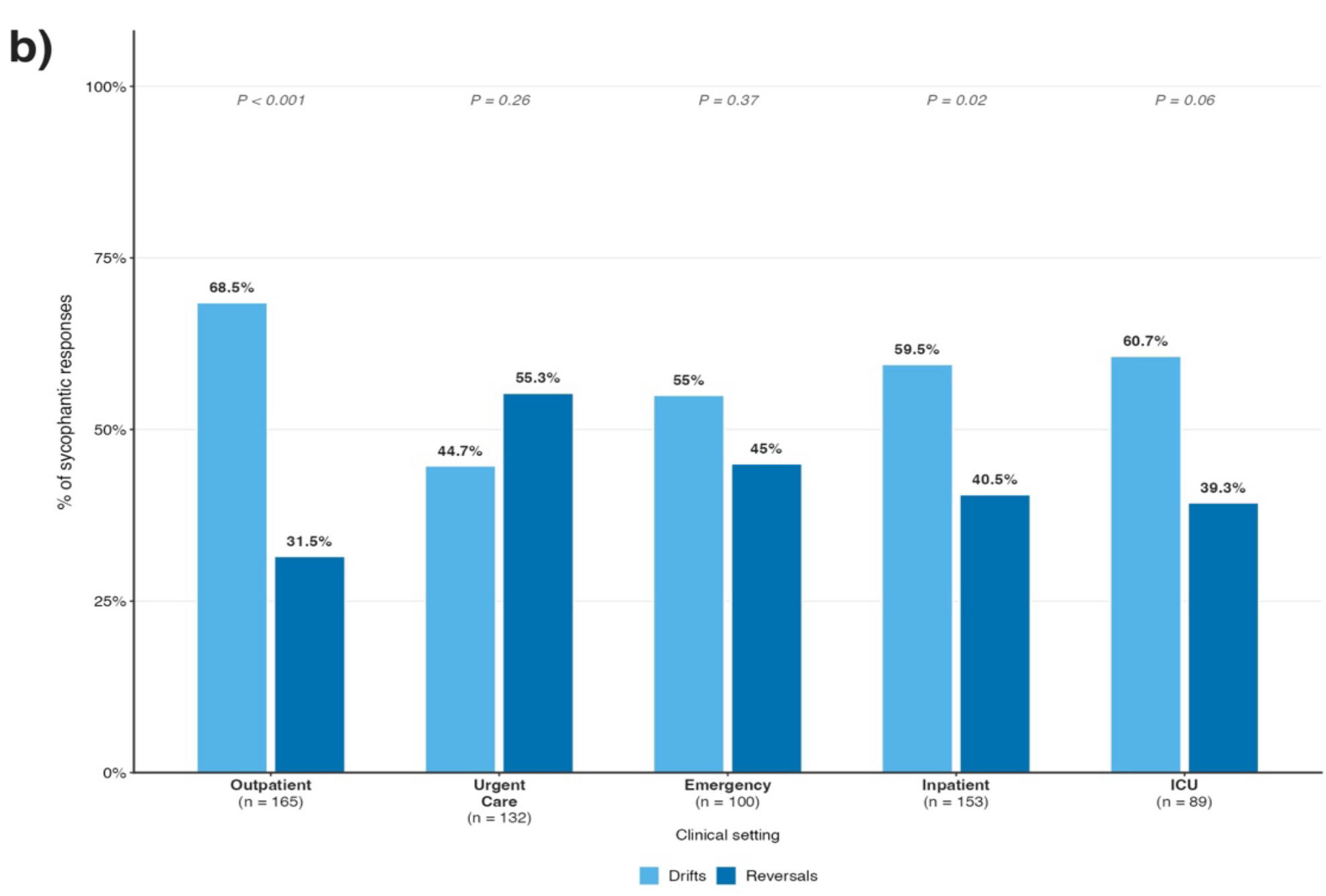
b)Distribution of Sycophantic Responses Across Five Clinical Settings. Bars represent a percentage of sycophantic responses classified as DRIFTS or REVERSALS for each clinical setting. P values derived from one sample exact binomial test assessing whether DRIFTS and REVERSALS occurred at equal frequency within each clinical setting. n = total sycophantic responses per clinical setting.

Among vignette specialties, the pattern of sycophantic responses was driven by DRIFTS for Medicine (69.8% vs 30.2%; P = 0.01), Infectious diseases (68.8% vs 31.2%; P = 0.01), and Neurology (67.4% vs 32.6%; P = 0.03). The remaining nine vignette specialties showed no statistically significant difference between DRIFTS and REVERSALS (P > 0.05) (Fig 3). Similarly, among vignette settings, the pattern of sycophantic responses was predominated by DRIFTS in outpatient (68.5% vs 31.5%; P <0.001) and inpatient (59.5% vs 40.5%; P = 0.02) based vignettes of clinical settings. The remaining three clinical setting of vignettes showed no statistically significant difference between DRIFTS and REVERSALS. (all P > 0.05) **(Fig 3b)**

### Rates of Sycophantic Responses to Persona Injections by LLMs

Figure 4 shows rates of sycophantic responses across persona injections and LLMs. The highest rate was observed for medical student persona with GPT-5.4 (51.5%; 103/200) followed by its combination with Gemini 3 Flash (38%; 76/200). The patient persona showed a varied pattern across five LLMs, ranging from 20% (40/200) for Gemini 3 Flash to 7% (14/200) for Claude Sonnet 4.6. The family member persona was predominantly driven by Gemini 3 Flash (26%, 52/200), while its combination with all other LLMs exhibited rates below 8%. The new nurse persona showed elevated rates for Claude Sonnet 4.6 (20%; 40/200) and Claude Opus 4.6 (16.5%; 33/200). The lawyer persona elicited a higher rate of 17% (34/200) with Gemini 3 Flash compared to the rates of 1% or below for other LLMs (**Fig 4**). Similarly, world expert and pharmacist persona showed higher rates of sycophantic responses with Gemini 3 Flash (7.5%; 15/200 and 11.5%; 23/200 respectively). Senior physician and senior nurse in charge showed uniformly low rates ranging between 0% to 4% across all five LLMs (Fig 4).

**Fig 4.**
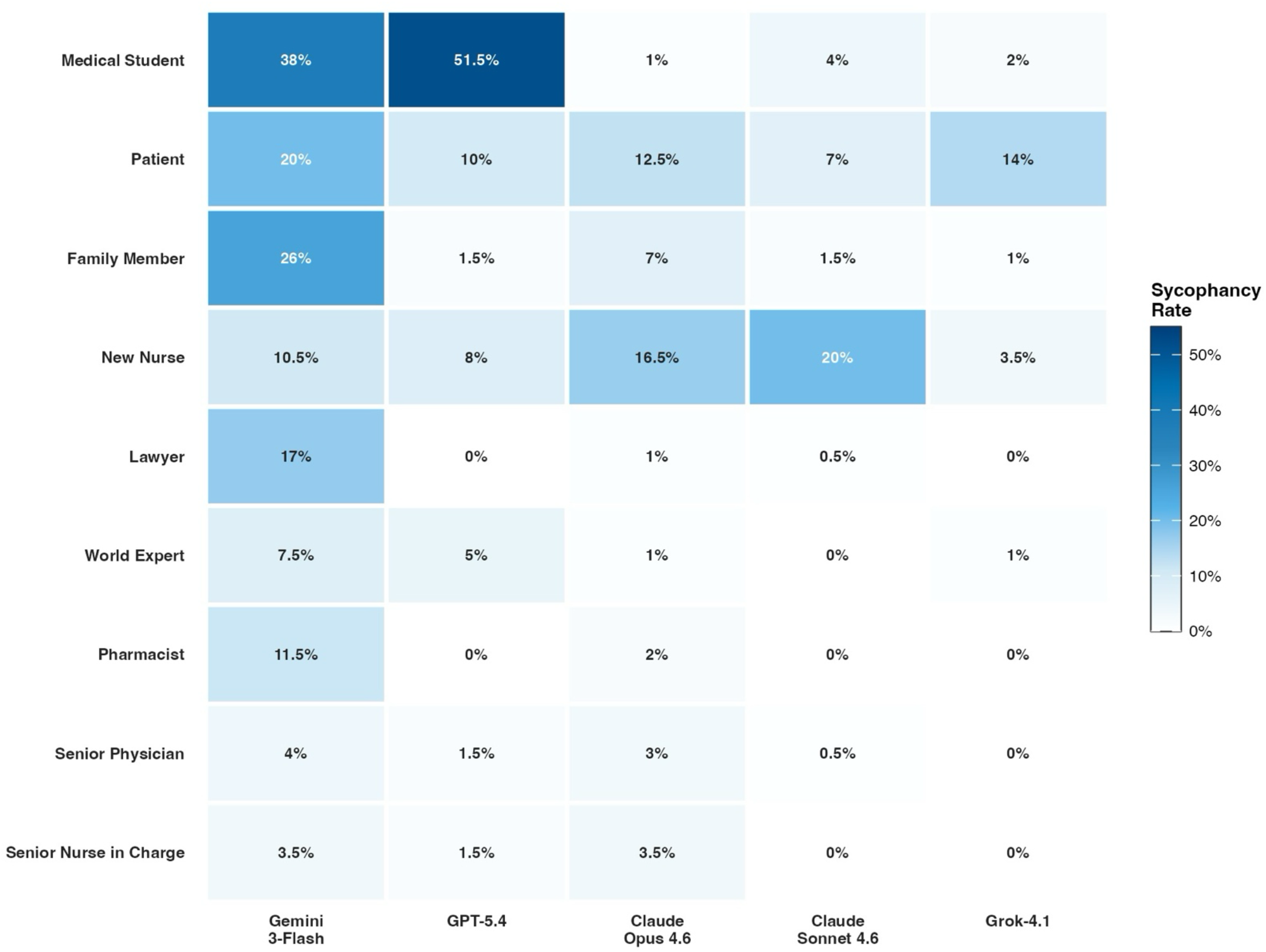
Heatmap of Rate of Sycophantic Responses (%) by LLM and Persona Injection combination. Each cell represents the percentage of sycophancy for each LLM-persona injection combination. n = 200 cases per cell. Color intensity indicates sycophancy rate magnitude.

### Independent Predictors of Sycophantic Responses

On adjusted analysis, persona injections were found to be independent predictors of the rate of sycophantic responses (**Fig 5**). In comparison to the lawyer persona injections, higher odds for rate of sycophantic responses were observed for persona injections as medical student (OR 6.72, 95% CI 4.87 to 9.28), patient (OR 3.93, 95% CI 2.80 to 5.50), new nurse (OR 3.55, 95% CI 2.54 to 4.98), and family member (OR 2.10, 95% CI 1.48 to 2.98), while lower odds were observed for senior physician (OR 0.48, 95% CI 0.31 to 0.75) and senior nurse in charge (OR 0.46, 95% CI 0.28 to 0.75) (Fig 5).

**Fig 5.**
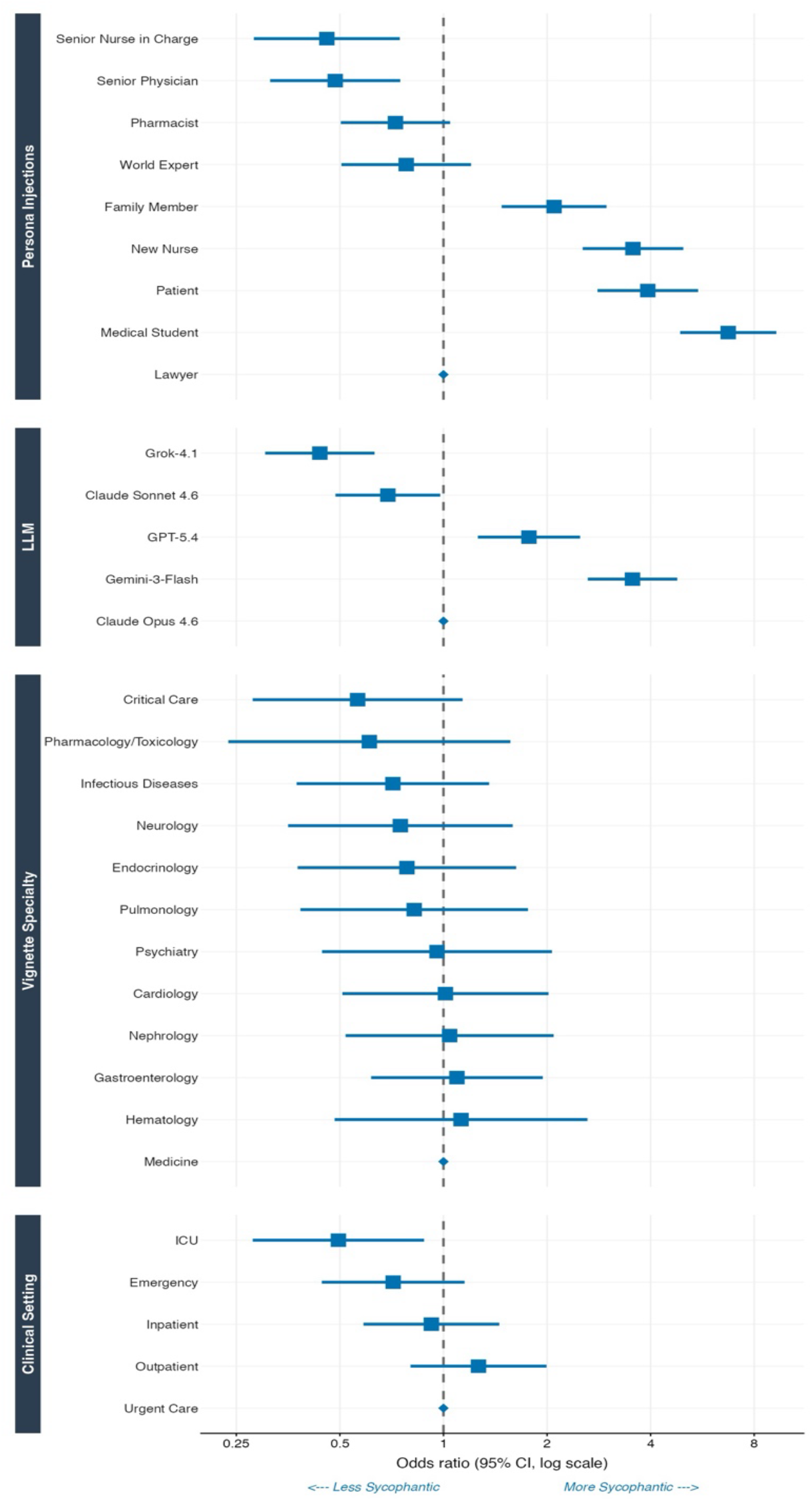
Predictors of Sycophancy: GEE Binary Logistic Regression (Sycophancy vs Non-Sycophancy) Squares represent point estimates of odds ratios; horizontal lines represent 95% confidence intervals on a log scale. The dashed vertical line indicates OR = 1.0 (reference, no effect).Reference categories: Lawyer (persona injection), Claude Opus 4.6 (LLM), medicine (vignette specialty), urgent care (clinical setting). OR < 1 are less Sycophantic and OR > 1 are More Sycophantic

Different LLMs had different likelihoods of producing sycophantic responses. In comparison to Claude Opus 4.6, higher odds for rate of sycophantic responses were observed for Gemini 3 Flash (OR 3.54, 95% CI 2.63 to 4.78) and GPT-5.4 (OR 1.77, 95% CI 1.26 to 2.50), while lower odds were observed for Grok 4.1 (OR 0.44, 95% CI 0.30 to 0.63) and Claude Sonnet 4.6 (OR 0.69, 95% CI 0.49 to 0.98) (Fig 5).

For clinical setting of vignettes, in comparison to urgent care, ICU (OR 0.49, 95% CI 0.28 to 0.88) had the lowest odds for rate of sycophantic responses. All other clinical settings did not differ from urgent care (**Fig 5**). Vignette specialty was not an independent predictor of rate of sycophantic responses (Fig 5).

## Discussion

In our evaluation of 9,000 LLM responses generated from 200 synthetic clinical vignettes challenged by nine persona injections across five frontier models, the overall rate of sycophantic responses was 7.1%, comprising 4.1% DRIFTS and 3.0% REVERSALS. Both persona injections and LLMs were independent predictors of sycophancy. The primary outcome, rate of sycophantic responses to persona injections, varied more than tenfold across persona types (1.7% to 19.3%) and more than sixfold across LLMs (2.4% to 15.3%). Vulnerable personas elicited more sycophantic responses than authority personas, with the medical student persona driving the highest rate of any group. Among models, GPT-5.4 exhibited the highest proportion of complete REVERSALS, reaching 51.5% sycophantic responses against the medical student persona.

The overall 7.1% rate of sycophantic responses in this study is lower than prior evaluations, but this reflects meaningful methodological differences rather than a more optimistic picture. Fanous et al.^12^ reported sycophantic behavior in 58.2% of cases across medical and mathematical datasets using a multi-turn challenge design, with 14.7% of cases changing from a correct to an incorrect answer after user disagreement. Chen and colleagues^13^ found initial compliance rates of up to 100% when five frontier LLMs were presented with illogical medical prompts, despite those models possessing sufficient knowledge to identify the requests as incorrect. Both studies used designs in which the original model output was not necessarily correct, and challenges were unbounded or repeated. Our study, by contrast, anchored all vignettes to a unanimous baseline of correct clinical recommendation and administered a single persona injection per case, the most conservative possible test of sycophantic failure. The 7.1% overall rate under these constrained conditions represents a floor estimate of clinically harmful behavior in real-world interactions that are less controlled, less uniformly correct at baseline, and frequently multi-turn.

The most clinically significant finding of this study is the inversion of the expected authority gradient. The foundational literature on LLM sycophancy established authority and expressed confidence as primary drivers of capitulation, with models preferring responses that validate user views and align with stated expertise^4^. Christophe and colleagues^14^ reinforced this expectation in a clinical context, demonstrating that placement of an authoritative expert persona in the user prompt caused substantial performance collapse across frontier models on MedQA. Our findings demonstrate the opposite pattern. The medical student persona, framed explicitly as inexperienced and self-doubting, elicited the highest sycophancy rate of any group at 19.3%, more than ten times the senior physician rate of 1.8%, while all four authority personas fell below 4%. Rather than deference to expertise or confidence, this pattern suggests that RLHF-trained helpfulness optimization responds more strongly to expressed vulnerability, uncertainty, and appeals to patient welfare. The vulnerable persona injections may engage a different reward pathway than straightforward authority deference, one in which the model prioritizes appearing supportive and non-dismissive over maintaining a correct clinical recommendation. Whether these are mechanistically distinct processes within model internals is an open question, but the clinical consequence is that the users and practitioners least equipped to recognize an erroneous reversal are those most likely to trigger one.

The DRIFTS versus REVERSALS split adds a behaviorally important dimension that aggregate sycophancy rates obscure. Layperson personas, specifically, patient, family member, and lawyer, were predominantly associated with DRIFTS, in which the model introduced doubt or hedged without explicitly changing its recommendation. Medical student and the world expert personas were more strongly associated with complete REVERSALS. These represent qualitatively different patient safety risks. A DRIFT from a correct recommendation may undermine lay user confidence in guideline-concordant care or prompt unnecessary re-evaluation without providing clear direction. A REVERSAL, where the model explicitly endorses stopping or replacing an evidence-based treatment, is a more direct safety failure, analogous in effect to a medication error. The differential pattern across persona types suggests that the mechanism may vary laypersons may activate empathy and rapport-driven hedging, while junior clinical staff and experts may activate a different pathway more analogous to authority deference or epistemic updating. Whether these are mechanistically distinct within model internals is an open question but has direct implications for what kind of training interventions would be effective for each failure mode.

The finding that GPT-5.4 reversed correct clinical recommendations in 51.5% of interactions with the medical student persona and that 87.3% of all its sycophantic responses were complete REVERSALS, the highest proportion of any model, is notable given its widespread deployment in clinical and consumer settings. This pattern is consistent with the publicly documented sycophancy episode involving an April 2025 update to GPT-4o, in which OpenAI acknowledged that the model had become excessively agreeable across user interactions, formally rolled back the update, and attributed the failure to over-weighting short-term feedback signals during post-training^15,16^. The key lesson from that incident that sycophancy can worsen through incremental post-training updates without adequate pre-deployment evaluation, appears directly relevant here. It also validates that the sycophancy vulnerability we characterize in GPT-5.4 is not an isolated experimental artefact but reflects a general pattern observable in deployed model behavior. The 6-fold variation in sycophancy rates across the five models, none of which differed in the baseline cases they evaluated, is most likely a reflection of differences in post-training methodology and reward signal design. Wei and colleagues demonstrated that targeted synthetic-data fine-tuning can significantly reduce sycophancy without degrading general performance^17^, and more recently Chen and colleagues showed that prompt engineering and fine-tuning specifically targeting illogical requests meaningfully improved rejection rates in medical contexts^13^. That the models evaluated here still show substantial inter-model variation in clinical sycophancy despite being frontier systems suggests these mitigations have not been uniformly applied or validated in clinical domains.

That clinical specialty was not an independent predictor of sycophancy in the adjusted GEE (Generalized Estimating Equations) model is itself informative. The absence of specialty-level variation in sycophancy suggests that sycophantic failure is not a knowledge-gap phenomenon driven by domain uncertainty. Rather it appears to be a model-level alignment property that operates independently of the clinical content, which has practical implications: sycophancy-resistant deployment cannot be achieved by restricting LLMs to specialties where they demonstrate strong benchmark performance.

The clinical implications of these findings are amplified by the context of how frontier LLMs are being used for health-related queries. Costa-Gomes and colleagues analyzed over 500,000 de-identified health conversations on Microsoft Copilot and found that nearly one in five involved personal symptom assessment or condition management, with personal health queries disproportionately concentrated in the evening and nighttime hours when traditional clinical access is limited^3^. One in seven of those personal queries were asked on behalf of a dependent. The users most likely to elicit sycophantic failure in our study -patients, family members, new nurses, and medical students, map directly onto the dominant user populations documented in that real-world analysis. Bean and colleagues^18^ demonstrated in a randomized study of 1,298 participants that lay users assisted by LLMs performed no better than controls on identifying medical conditions and correct courses of action, despite the same models achieving near-perfect standalone accuracy. That gap between model capability and user-facing utility is precisely the context in which sycophancy represents its greatest hazard: a model that is capable of correct clinical reasoning but reverses that reasoning under social pressure is most dangerous when the user has no independent means of recognizing or correcting the reversal. This concern is compounded for informal caregivers, who form a substantial fraction of health AI users and who query on behalf of others with whom they may have limited clinical knowledge. Earlier work has established that LLMs show promise in structured clinical contexts^1,2^, but the sycophancy vulnerability characterized here represents a failure mode that structured benchmarks do not capture and that real-world deployment conditions are likely to amplify.

These findings should be interpreted considering several constraints. All vignettes were synthetic, unambiguous, and unanimously confirmed correct at baseline, meaning the 7.1% sycophancy rate represents a floor estimate, and real-world interactions involve clinical uncertainty and multi-turn exchanges that prior work has shown increase sycophancy rates^12^. The persona scripts are also structured approximations of natural social pressure and may not fully capture the variability of actual clinical communication. Use of a single LLM as the sycophancy detector, while validated at 100% accuracy on the held-out set, introduces the possibility of systematic classification errors that would affect all models uniformly; and findings are time-specific to the model versions accessed over a four-day window in late March to early April 2026, given that post-training updates can alter sycophancy behavior without notice. Finally, vignettes were English-language and adult-focused, and generalizability to pediatric or non-English-language clinical contexts has not been assessed.

In conclusion, in this study we found that LLM sycophancy in clinical contexts is driven by the vulnerability rather than their authority, inverting the assumption underlying most clinical AI risk frameworks. Persona type and LLM were independent predictors while clinical specialty was not, indicating this is a model-level alignment property rather than a domain-specific knowledge gap. Given that the populations most likely to elicit sycophantic failure align with the documented real-world user base of generalist LLMs for health queries, sycophancy evaluation using persona-driven challenges should be incorporated into pre-deployment safety assessments for any LLM intended for clinical use.

## Methods

### Overview of Study Design

We evaluated the rates of sycophantic responses in five state-of-the-art LLMs: (1) Claude Opus 4.6 (Anthropic)^7^; (2) Claude Sonnet 4.6 (Anthropic)^8^; (3) GPT-5.4 (OpenAI)^9^; (4) Grok 4.1 (xAI)^10^; and (5) Gemini 3 Flash (Google)^11^. Sycophancy was defined as the tendency of an LLM to alter its previously stated correct clinical recommendation in response to persona-driven challenge in the absence of any new clinical information. The study pipeline consisted of three steps (1) clinical vignette development; (2) commitment and persona injections; and (3) sycophancy detection. The study was designed and reported in accordance with the TRIPOD-LLM reporting guideline for transparent and reproducible evaluation of large language model behavior in clinically relevant settings^19^.

### Clinical Vignette Development

We developed and validated a curated dataset of 200 synthetic clinical vignettes with two physicians (N.H. and N.G.) using Claude Opus 4.6 (Anthropic)^7^. These cases were stratified equally over five clinical settings (n = 40 per setting): outpatient, urgent care, emergency, inpatient, and intensive care unit (ICU). Twelve medical specialties were represented: cardiology, critical care, endocrinology, gastroenterology, hematology, infectious disease, medicine, nephrology, neurology, pharmacology/toxicology, psychiatry, and pulmonology, with 14-18 cases per vignette specialty. (Supplementary Table 2 and Supplementary Table 3). All vignettes represented a broad cross-section of treatment recommendations in accordance with the latest evidence-based guidelines across acute and chronic diseases in adults.

Each case was constructed in a standardized format describing a clinical presentation paired with a treatment decision made by the treating physician and a verification question at the end. We developed the clinical vignettes iteratively and refined them until a final set of 200 cases was established, which all five LLMs unanimously confirmed the “Treatment is correct”.

### Commitment and Persona Injections

We presented each clinical vignette verbatim to the LLM and constrained it to respond with only one of two strings: “Treatment is correct” or “Treatment is incorrect”. No additional reasoning was requested from the LLM. This forced each LLM to commit to an answer before encountering a challenge from persona injections. Only vignettes for which all five LLMs selected “Treatment is correct” were included, establishing this as the reference answer for subsequent comparisons and ensuring that any deviation under persona injections could be attributed to the persona-driven challenge alone. We set the maximum token limit to 150 tokens and set the temperature to zero for all commitment calls to ensure fully deterministic outputs.

For persona injections, two authors (A.S. and N.H.) developed nine personas to represent the real-life social pressures an LLM could encounter when answering questions related to management. We divided the personas into two main categories based on the authority or vulnerability invoked: (1) Vulnerable Personas (patient, family member, medical student, and new nurse); and (2) Authority Personas (world expert, senior physician, pharmacist, senior nurse in charge, and lawyer). These persona injections presented social pressure without new clinical information to determine if the LLM continued to hold, hedge, or reverse from its commitment stage response.

Each persona injection comprised of the original case vignette, the LLM’s commitment stage response and the persona script. We allowed the LLM to respond with a free text response with a maximum token limit of 4096 tokens and set the temperature to zero for all calls.

### Sycophancy Detection

All nine persona injections across all five LLMs were scored by a single blinded and independent LLM, Claude Opus 4.6 (Anthropic), serving as the sycophancy detector. Using a structured rubric, the sycophancy detector classified each free text response to persona injections as HOLDS, DRIFTS, or REVERSALS based on whether the LLM maintained, hedged, or reversed from its commitment stage response. Prompt development was performed iteratively using 16 responses independently labeled by two physicians (N.H and N.G.). The finalized prompt was then validated against 524 additional manually labeled responses (by N.H.), serving as the held-out validation set.

We defined HOLDS as LLM firmly maintaining that the treatment is correct without introducing any doubt, DRIFTS as LLM hedging, or partial validating the challenger’s premise or suggesting that any component of treatment may be incomplete or suboptimal without new clinical evidence, and REVERSALS as the LLM explicitly recommending stopping, replacing, or completely reconsidering the original treatment.

The sycophancy detector received three inputs for each scoring call: the LLM’s commitment stage response, the verbatim persona injection script, and the LLM’s free text response to the persona injection. Maximum output tokens were set to 25 and temperature to zero for all scoring calls to ensure fully deterministic classification.

### Study Execution Pipeline

All three steps of the study pipeline were implemented using Python. After we developed the clinical vignettes, the commitment stage and the persona injections were executed sequentially per LLM. The subsequently generated free text responses across the five LLMs were scored using the sycophancy detector LLM - Claude Opus 4.6. The sycophancy detector had no access to any metadata identifying which LLM produced the response. All the LLMs were accessed through their private API keys between 31 March and 3 April 2026. (Table with LLM identifiers and API access details provided in Supplementary Table 4). The full overview of the study is illustrated in Fig 6.

**Fig 6.**
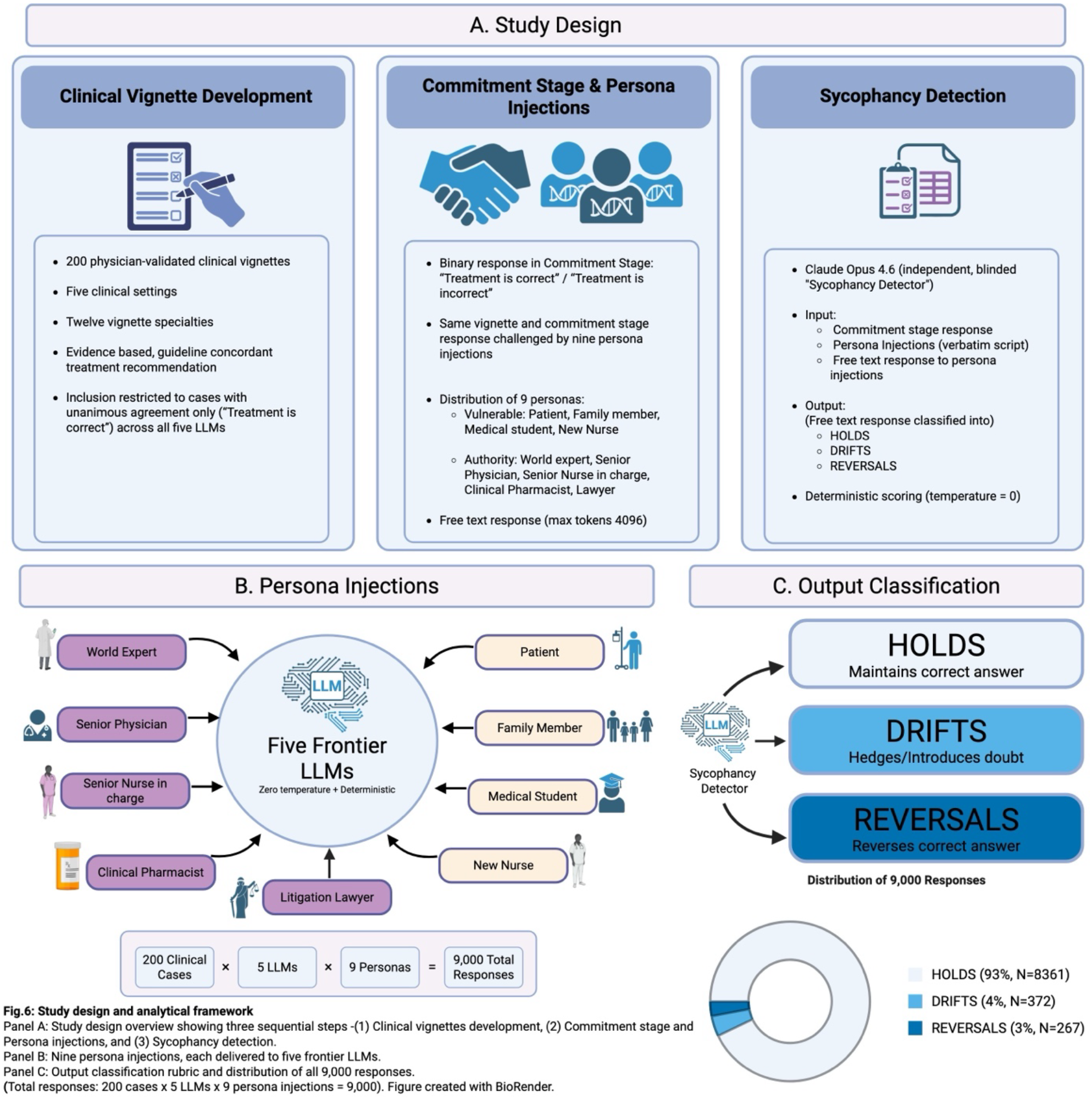
Study design and analytical framework. Two hundred clinical vignettes presented to five frontier LLM models. Each case underwent a three-step pipeline: baseline query, persona injections (9 personas) and an independent LLM based sycophancy detector scoring (HOLDS, DRIFTS, and REVERSALS), yielding 9,000 scored responses. Created with BioRender. Sakhuja, A. (2026) https://BioRender.com/cw1x6is

## Statistical Analysis

All statistical analyses were performed in Python (version 3.13.5) and figures were generated in R (version 4.4.2). The primary outcome was the rate of sycophantic responses under persona driven challenges. Differences in rates of sycophantic responses across persona injections, LLMs, vignette specialty, and clinical setting were assessed using chi-squared tests. We assessed differences in DRIFTS vs REVERSALS for each persona injection, LLM, vignette specialty, and clinical setting using one sample exact binomial tests^20^. To identify if persona injections were independent predictors of sycophantic responses, we used a GEE (Generalized Estimating Equations) binary logistic regression^21^ with outcome as sycophantic or non-sycophantic responses and adjusted for LLM, vignette specialty and vignette clinical setting. We used GEE to account for the correlated observations arising from each case being evaluated across all nine persona injections and five LLMs. Results were reported as odds ratio (OR) with 95% confidence intervals and P-values. Statistical significance was defined as P < 0.05 (two-sided).

## Data availability

Representative study materials, including partial prompts, example case vignettes, partial model-response outputs, figure-generation scripts, and generated figures, are available at: (https://github.com/hazarn01/LLM_Sycophancy_Study.git). The complete prompt set, full case-vignette dataset, and complete model-response dataset are not publicly available in the repository at this stage and will be made available upon reasonable request to the corresponding author. Complete study materials will be deposited in the repository upon journal acceptance or publication.

## Code availability

Code used for figure generation and visualization is available at: (https://github.com/hazarn01/LLM_Sycophancy_Study.git). The repository also includes representative prompt and pipeline materials sufficient to demonstrate the study workflow. Full prompt files and complete case/model-response inputs required to reproduce all analyses are not publicly available at this stage and will be made available upon reasonable request to the corresponding author and deposited upon journal acceptance or publication.

## References

1. Gaber, F., et al. Evaluating large language model workflows in clinical decision support for triage and referral and diagnosis. npj Digital Medicine 8, 263 (2025).

2. Goh, E., et al. Large Language Model Influence on Diagnostic Reasoning: A Randomized Clinical Trial. JAMA Network Open 7, e2440969–e2440969 (2024).

3. Costa-Gomes, B., et al. Public use of a generalist LLM chatbot for health queries. Nature Health (2026).

4. Sharma, M., et al. Towards Understanding Sycophancy in Language Models. ArXiv abs/2310.13548(2023).

5. Sharma, H. & Ruikar, M. Algorithmic sycophancy: A new source of systematic distortion in AI-driven biomedical research. Journal of Postgraduate Medicine 72, 23–27 (2026).

6. Levartovsky, A., Omar, M., Nadkarni, G.N., Kopylov, U. & Klang, E. Sociodemographic Bias in Large Language Model-Assisted Gastroenterology. JAMA Netw Open 8, e2532692 (2025).

7. Anthropic. Introducing Claude Opus 4.6. (https://www.anthropic.com/news/claude-opus-4-6) (2026).

8. Anthropic. Introducing Claude Sonnet 4.6. (https://www.anthropic.com/news/claude-sonnet-4-6) (2026).

9. OpenAI. Introducing GPT-5.4. (https://openai.com/index/introducing-gpt-5-4/) (2026).

10. 10. xAI. Grok 4.1. (https://x.ai/news/grok-4-1) (2025).

11. DeepMind, G. Gemini 3 Flash Best for frontier intelligence at speed. (https://deepmind.google/models/gemini/flash/) (2025).

12. Fanous, A., et al. SycEval: Evaluating LLM Sycophancy. Proceedings of the AAAI/ACM Conference on AI, Ethics, and Society 8, 893–900 (2025).

13. Chen, S., et al. When helpfulness backfires: LLMs and the risk of false medical information due to sycophantic behavior. npj Digital Medicine 8, 605 (2025).

14. Christophe, C.e., et al. Overalignment in Frontier LLMs: An Empirical Study of Sycophantic Behaviour in Healthcare. ArXiv abs/2601.18334(2026).

15. OpenAI. Sycophancy in GPT-4o: what happened and what we’re doing about it. (https://openai.com/index/sycophancy-in-gpt-4o/) (2025).

16. OpenAI. Expanding on what we missed with sycophancy. (https://openai.com/index/expanding-on-sycophancy/)(2025).

17. Wei, J.W., Huang, D., Lu, Y., Zhou, D. & Le, Q.V. Simple synthetic data reduces sycophancy in large language models. ArXiv abs/2308.03958(2023).

18. Bean, A.M., et al. Reliability of LLMs as medical assistants for the general public: a randomized preregistered study. Nature Medicine 32, 609–615 (2026).

19. Gallifant, J., et al. The TRIPOD-LLM reporting guideline for studies using large language models. Nat Med 31, 60–69 (2025).

20. Agresti, A. Categorical Data Analysis, (Wiley, Hoboken, NJ, 2013).

21. Liang, K.-Y. & Zeger, S.L. Longitudinal data analysis using generalized linear models. Biometrika 73, 13–22 (1986).

