## Supplementary Information for "Evaluating Sycophancy in Frontier Models Using Persona-Driven Challenge"

**Supplementary Tables**

**Supplementary Table 1 | Table with Rates of Sycophantic responses for twelve Vignette Specialties**

| **Discipline** | **Total responses** | **DRIFTS (%)** | **REVERSALS (%)** | **Sycophancy Rate (%)** |
| --- | --- | --- | --- | --- |
| Hematology | 765 | 34 (4.4) | 41 (5.4) | 75 (9.8) |
| Gastroenterology | 765 | 38 (5.0) | 33 (4.3) | 71 (9.3) |
| Psychiatry | 675 | 29 (4.3) | 23 (3.4) | 52 (7.7) |
| Nephrology | 765 | 33 (4.3) | 23 (3.0) | 56 (7.3) |
| Cardiology | 810 | 33 (4.1) | 25 (3.1) | 58 (7.2) |
| Pharmacology/Toxicology | 630 | 27 (4.3) | 17 (2.7) | 44 (7.0) |
| Medicine | 765 | 37 (4.8) | 16 (2.1) | 53 (6.9) |
| Pulmonology | 810 | 30 (3.7) | 24 (3.0) | 54 (6.7) |
| Infectious Diseases | 765 | 33 (4.3) | 15 (2.0) | 48 (6.3) |
| Endocrinology | 765 | 28 (3.7) | 19 (2.5) | 47 (6.1) |
| Neurology | 765 | 31 (4.1) | 15 (2.0) | 46 (6.0) |
| Critical Care | 720 | 19 (2.6) | 16 (2.2) | 35 (4.9) |
| **TOTAL** | **9000** | **372** | **267** | **639 (7.1)** |

Frequency of sycophantic behavior across 12 medical specialties. Model responses to persona challenges were classified as Drifts (hedging/equivocating) or Reversals (completely changing the initial correct management plan). The overall Sycophancy Rate represents the combined proportion of Drifts and Reversals. Data aggregates 9,000 total challenges across 5 LLMs.

**Supplementary Table 2 | Table with distribution of cases by clinical setting**

| **Clinical Setting** | **Cases** |
| --- | --- |
| Urgent Care | 40 |
| Emergency | 40 |
| Inpatient | 40 |
| Intensive Care Unit (ICU) | 40 |
| Outpatient | 40 |
| **TOTAL** | **200** |

Breakdown of the 200 physician-curated clinical vignettes utilized in the study, stratified equally across five distinct clinical settings.

**Supplementary Table 3 | Table with distribution of cases by vignette specialty**

| **Vignette Specialty** | **Cases** |
| --- | --- |
| Cardiology | 18 |
| Critical Care | 18 |
| Endocrinology | 17 |
| Gastroenterology | 17 |
| Hematology | 17 |
| Infectious Disease | 17 |
| Medicine | 17 |
| Nephrology | 17 |
| Neurology | 17 |
| Pulmonology | 16 |
| Psychiatry | 15 |
| Pharmacology/Toxicology | 14 |
| **TOTAL** | **200** |

Breakdown of the 200 physician-curated clinical vignettes utilized in the study, categorized across 12 distinct medical specialties.

**Supplementary Table 4 | Table of LLMs identifiers and access details**

| **LLMs** | **Developer** | **LLM string** | **Access method** |
| --- | --- | --- | --- |
| Claude Opus 4.6 | Anthropic | claude-opus-4-6 | Private API key |
| Claude Sonnet 4.6 | Anthropic | claude-sonnet-4-6 | Private API key |
| GPT-5.4 | OpenAI | gpt-5.4 | Private API key |
| Grok 4.1 | xAI | grok-4-1-fast-non-reasoning | Private API key |
| Gemini 3 Flash | Google | gemini-3-flash-preview | Private API key |

Details of the five commercial Large Language Models evaluated in the study, including their developers, the specific model version strings utilized for API queries, and the method of access. All models were queried securely via private API keys.

**Supplementary Table 5 | Table with Chi-square**

| **Test** | **Predictor** | **P-value** |
| --- | --- | --- |
| Chi-square | LLM | < 0.001 |
| Chi-square | Persona Injections | < 0.001 |
| Chi-square | Vignette Specialty | 0.018 |
| Chi-square | Clinical Setting | < 0.001 |

Statistical significance was defined as P < 0.05
